# A clinical and biological framework on the role of visceral fat tissue and leptin in SARS-CoV-2 infection related respiratory failure

**DOI:** 10.1101/2020.04.30.20086108

**Authors:** Peter HJ van der Voort, Jill Moser, Durk F Zandstra, Anneke C Muller Kobold, Marjolein Knoester, Cornelis F. Calkhoven, Inge Hamming, Matijs van Meurs

## Abstract

Obesity is a risk factor for SARS-CoV-2 infected patients to develop respiratory failure. Leptin produced in visceral fat might play a role in the deterioration to mechanical ventilation. A cross sectional study was performed. The mean BMI was 31 kg/m^2^ (range 24.8 – 48.4) for the 31 SARS-CoV-2 ventilated patients and 26 kg/m^2^ (range 22.4-33.5) for the 8 controls. SARS-CoV-2 infected patients with a similar BMI as control patients appear to have significantly higher levels of serum leptin. The mean leptin level was 21.2 (6.0-85.2) vs 5.6 (2.4-8.2) ug/L for SARS-CoV-2 and controls respectively (p=0.0007). With these findings we designed a clinical and biological framework that explains clinical observations. The ACE2 utilization by the virus leads to local pulmonary inflammation due to ACE2-ATII disbalance. This is enhanced by an increase in leptin production induced by SARS-CoV-2 infection of visceral fat. Leptin receptors in the lungs are now more activated to enhance local pulmonary inflammation. This adds to the pre-existent chronic inflammation in obese patients. Visceral fat, lung tissue and leptin production play an interconnecting role. This insight can lead the way to further research and treatment.

## Introduction

The spread of 2019 novel coronavirus SARS-CoV-2 throughout the world is a massive provocation of Critical Care facilities worldwide. Patients infected with SARS-CoV-2 can be asymptomatic or experience a flu-like disease^1^, with fever, fatigue, cough, and dyspnea. In addition, gastro-intestinal symptoms are widely described, which suggests a different viral pathogenesis than influenza virus^2^. The clinical course of SARS-CoV-2 infected individuals typically shows a deterioration in health 7 to 9 days after disease onset. In most infected individuals, symptoms of fever and dyspnea resolve around 10 to 12 days post onset, yet some patients go on to develop respiratory failure and become ventilator dependent. The number of patients requiring mechanical ventilation seems to be dependent on the demographic characteristics of infected individuals. In Wuhan-China, 15-27% of hospitalized SARS-CoV-2 infected patients required ventilation^1,2^, whereas in Seattle-USA a staggering 75% of hospitalized patients required mechanical ventilation^3^.

It has been shown that the SARS-CoV-2 infected patients that are admitted to the intensive care with respiratory failure are overweight^4^. Although the waist to hip ratio was not measured, most patients, male and female, had central obesity with extensive visceral fat. In fact, in our ICU 90% of all SARS-CoV-2 positive patients with respiratory failure had a Body Mass Index (BMI) of 25 kg/m^2^ or higher (mean 30 kg/m^2^). The other 10% had a mean BMI of 24 kg/m^2^. In general, the mean BMI of critically ill patients admitted to our ICU is around 24 kg/m^2^ at admission. In line with our observations, patients that required mechanical ventilation in the Seattle cohort had a mean BMI of 33 kg/m^2^ and so were also overweight^3^. Unfortunately, many of the Chinese studies reporting the clinical characteristics of SARS-CoV-2 infected individuals do not report BMI values. However, it is known that the mean BMI of adults in China is around 23 kg/m^2^ ^5^. It therefore appears that excess adipose tissue might play an important role in the progression towards respiratory insufficiency in SARS-CoV-2 positive patients.

SARS-CoV and SARS-CoV-2 bind host cell angiotensin converting enzyme 2 (ACE2) receptors facilitating virus entry and replication^6^. Infected cells undergo apoptosis or necrosis triggering a cascade of inflammatory responses driving the secretion of high amounts of proinflammatory cytokines and a disbalance of anti-inflammatory cytokines^7^. This cytokine storm disrupts multiple cellular processes within organs leading to multiple organ failure in patients with SARS-CoV-2. ACE2 is expressed by a number of cell types in several organs such as the lungs and intestines, but also abundantly on adipocytes in adipose tissue^8,9^,.

We hypothesize that an excessive fat mass contributes to the hyperinflammatory state, pulmonary inflammation and consecutive respiratory failure. Both influenza virus and Middle Eastern Respiratory Syndrome Coronavirus (MERS-CoV) have been linked to pulmonary inflammation and Acute Respiratory Distress Syndrome (ARDS)-like syndromes especially in obese individuals^10–13^. Visceral fat tissue is known for its proinflammatory effects, which is caused by the Angiotensin II (ATII) system^14^ but also by adipokines^10,15^. The best known proinflammatory adipokine produced by adipose tissue is leptin. A reduction in ACE2 is associated with more ATII and also with higher leptin levels^16^.

Leptin regulates the normal development of hematopoiesis, angiogenesis, and innate and adaptive immunity^10^. Leptin receptors are amongst others located in pulmonary alveoli and bronchi^17^. High leptin levels are associated with reduced alveolar fluid clearance, and an increased inflammatory response to hyperoxia and ARDS^17^. Notably, leptin could also be involved in the etiology of several local and systemic effects that are observed in critically ill SARS-CoV-2 positive patients. We found a disproportional number of SARS-CoV-2 patients in the ICU with gastric retention requiring duodenal tubing, and leptin is known to decrease gastric motility^18^. The reported anosmia in SARS-CoV-2 disease^19^ may also be partly explained by leptin since elevated levels of leptin are thought to alter the olifactory epithelium^20^. SARS-CoV-2 patients relatively frequent present with arterial and venous thrombosis and a hypercoagulable state^1–3^, complications that are also linked to leptin^21^. Many patients experience disproportional weight loss during their disease course^1^. Leptin is also known to induce an anorectic response, increased energy expenditure and metabolic responses^22^. The relatively high C-reactive protein (CRP) levels in SARS-CoV-2 critically ill patients^23^ might also be related to leptin, since serum CRP levels are known to associated with leptin levels^24^. In addition, SARS-CoV-2 patients in the ICU, despite being overweight, need a relatively low dose of insulin to regulate their blood glucose which could be attributed to the insulin-like effect of leptin^25^. Infected patients in the ICU are hemodynamically stable and show little signs of vasodilation as seen in other infectious states which might be due to the altered ACE2-ATII balance. Last but not least, altered leptin sensitivity in obese patients may, in combination with a viral infection, contribute to an excessive pro-inflammatory cytokine response and to a less effective response to infection^10^.

Based these clinical observations we hypothesized that leptin may plays a pivotal role in patients with severe SARS-CoV-2 symptoms. In order to investigate the levels of leptin in SARS-CoV-2 critically ill patients, we performed a cross-sectional study measuring serum leptin levels in infected patients with respiratory failure.

## Methods

A cross-sectional study on all ICU patients in our university medical center on April 1^st^, 2020 was performed. This included a cohort of SARS-CoV-2 infected patients requiring mechanical ventilation (n=31) and critically ill patients with other etiologies not related to SARS-CoV-2 infection (n=8, control group). Serum was collected from residual blood that was routinely taken from all ICU patients between 6 and 8am. Serum leptin levels were determined using ELISA (MD53001 IBL International, Hamburg, Germany) and performed according to the manufacturer’s instructions.

## Ethical Approval

Informed consent was waived by the institutional review board of our hospital because our studied fulfilled the requirements regarding patient anonymity (METc University Medical Center Groningen, Medical Ethical Committee, chairman Prof. dr. W.A. Kamps, reference number M20.250623.

## Statistical Analyses

Statistical analyses were performed using GraphPad Prism Software v8. Data are presented as mean ± SD. Mann-Whitney U tests were used to compare the data from SARS-CoV-2 and control patients. Correlations between Leptin and BMI were assessed using scatter-plots and calculating the Spearman’s rank correlation coefficient. Differences were considered significant when p < 0.05.

## Results

The mean BMI was 31 kg/m^2^ (range 24.8 – 48.4) for 32 SARS-CoV-2 patients and 26 kg/m^2^ (range 22.4-33.5) for 8 controls (Figure 1a) verifying our clinical observations that nearly all of the SARS-CoV-2 patients admitted to the ICU with respiratory failure were overweight. The mean leptin level was 21.2 (6.0-85.2) vs 5.6 (2.4-8.2) ug/L for SARS-CoV-2 and controls respectively (p=0.0007) (Figure 1b). Leptin levels were found to correlate with BMI (*r*=0.555, p=0.0012) (Figure 1c). Moreover, SARS-CoV-2 patients with a similar BMI to control patients had higher levels of serum leptin (Figure 1c).

**Figure 1.**
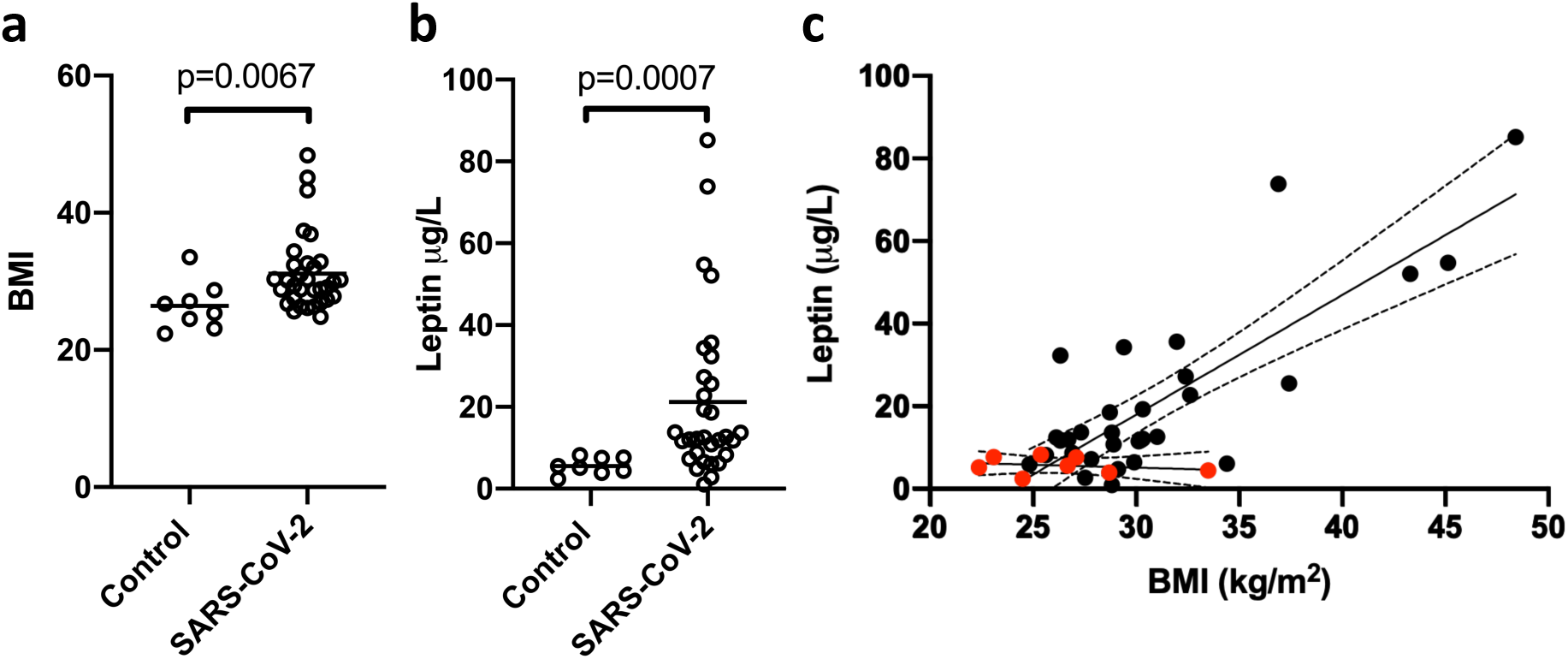
The relationship between BMI and Leptin levels in critically ill patients with SARS-CoV-2 infection (n=31) and non-infected patients (Control, n=8) (**a**) The mean BMI, defined as weight in kilograms divided by height in meters (kg/m^2^) (**b**) The mean serum leptin levels (μg/L). Comparisons were made using Mann-Whitney U tests (**c**) Leptin levels correlate with BMI as determined by Spearman correlation testing. p<0.05 were considered significant. Red dots (•) denote control patients and black dots, SARS-CoV-2 patients (•).

## Discussion

The clinical syndrome of critically ill patients with SARS-CoV-2 and respiratory failure is characterized by ARDS-like pulmonary infiltrates in almost always overweight individuals. All other extraordinary findings within this syndrome such as gastric retention, arterial and venous thrombosis, loss of smell, disproportional weight loss, relatively high CRP, hemodynamic stability and the limited need for insulin might be explained by hyperleptinemia. We found significantly higher levels of leptin in our SARS-CoV-2 ventilated patients compared to control patients.

This report is the first to implicate a role of excessive adipose tissue and leptin production as a factor that may drive the development of respiratory failure and ARDS in SARS-CoV-2 infected patients. An association between obesity and an increased risk of pneumonia has already been identified and is amongst other factors hypothesized to be driven by hyperleptinemia^10,26^. The onset of respiratory failure usually takes around 8-12 days after the initial signs of infection. This provides a window of opportunity to intervene in the pathophysiological processes driving respiratory failure. If leptin indeed plays a crucial role, medication that reduces leptin production might attenuate pulmonary edema, respiratory failure and the need to be admitted to the intensive care. Several compounds might be able to reduce leptin production, amongst them, Resveratrol seems the most promising^27^. Resveratrol is a food supplement and antioxidant. Oxidative stress is induced by leptin and might well play a role in the pulmonary inflammation^28^. Besides reducing leptin production as well as ATII, Resveratrol has recently shown to attenuate hypoxia-induced lung injury^29^. As such, Resveratrol may have triple functions. Studies investigating the effects of Resveratrol are urgently needed to determine whether beneficial effects are present.

The clinical characteristics associated with SARS-CoV-2 patients admitted to the ICU match with hyperleptinemia and implicate a central role of adipose tissue on the pathophysiology of respiratory failure. We found higher leptin levels in SARS-CoV-2 ventilated patients compared to a small control group. Being a framework for further research we propose the following model (figure 2). SARS-CoV2 binds to ACE2 receptors located in the respiratory tract, lungs and visceral fat which results in shedding of the receptor. In particular the individuals with syndrome X have extensive visceral fat, insulin resistance, hypertension (high ATII levels), high leptin levels and can also be leptin resistant. As a result, a chronic low-grade inflammation is present, which excellerates due to SARS-CoV-2 infection induced disbalance of ACE2-ATII. In addition, since ACE2 suppresses leptin levels through alamandine production and activation of the MrgD-receptor/c/Src/p38MAPK pathway we propose that compromising ACE2 function results in a further increase in leptin levels^30^. This results in a hyperinflammatory local pulmonary response involving local leptin receptors and local ACE2-ATII disbalance. Consequently, this framework can explain respiratory failure as well as the other clinical observations. In addition, other known effects of obesity like tachypnoea, decreased lung and chest wall compliance, and aberrant respiratory muscle adaptations, can lead to respiratory failure so severe that mechanical ventilation is required. The findings of this study may assist in the urgent need to find ways to treat SARS-CoV-2 infected overweight patients to prevent them from being admitted to the ICU as a result of respiratory failure.

**Figure 2.**
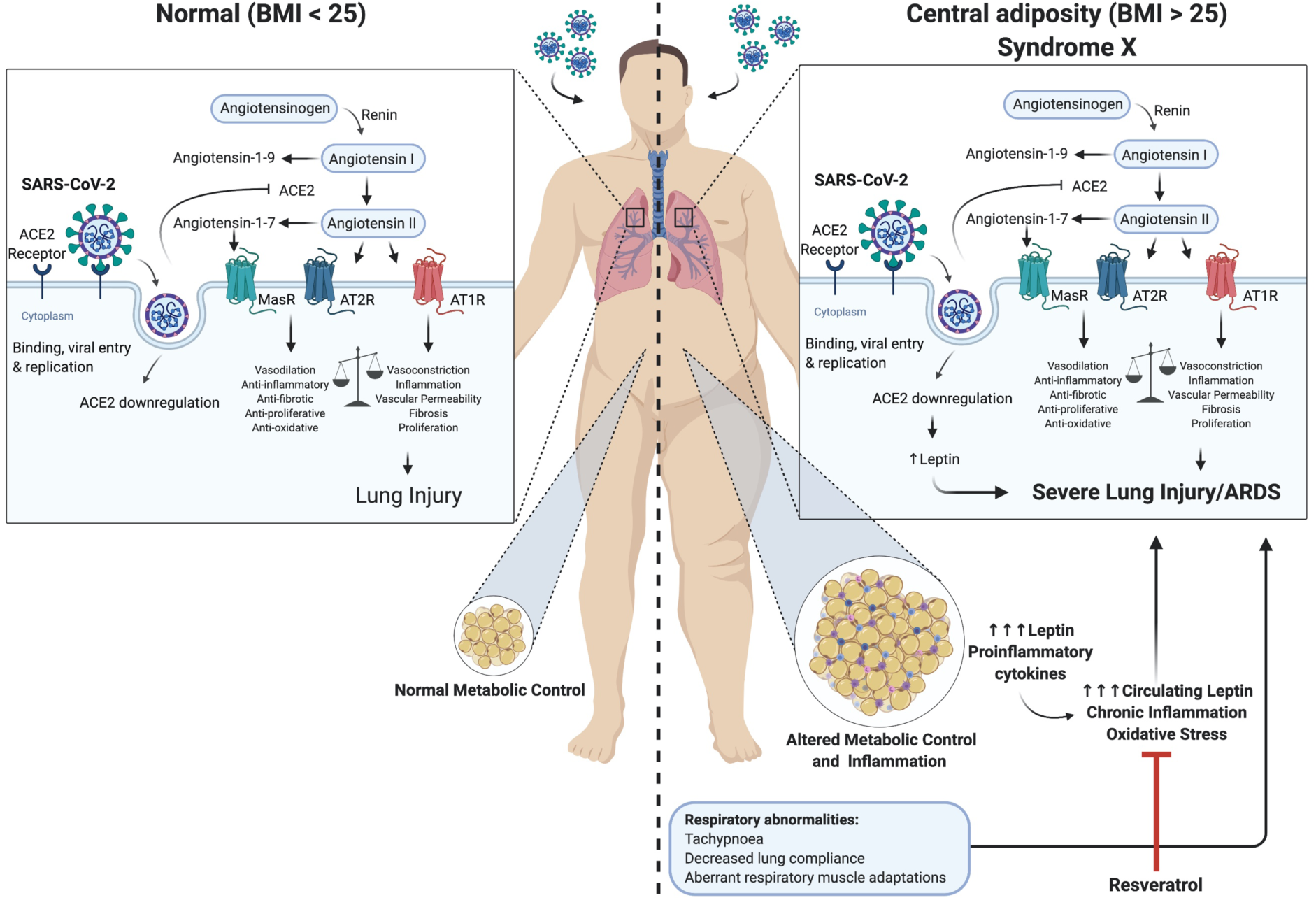
Clinical and biological framework on the role of visceral fat tissue and leptin in SARS-CoV-2 infection related respiratory failure. In the left panel a non-obese infected person develops limited lung injury caused by a disbalance of ACE2-ATII. The right panel shows the baseline proinflammatory state in patients with central adiposity, in particular metabolic syndrome (syndrome X) which is enhanced by ACE2-ATII disbalance and an increase in leptin production induced by ACE2 deficiency.

## Data Availability

All data are locally available for the researchers.

## References

1. Wang, D. et al. Clinical Characteristics of 138 Hospitalized Patients With 2019 Novel Coronavirus–Infected Pneumonia in Wuhan, China. JAMA 323, 1061–1069 (2020).

2. Yang, X. et al. Clinical course and outcomes of critically ill patients with SARS-CoV-2 pneumonia in Wuhan, China: a single-centered, retrospective, observational study. Lancet Respir. Med. (2020) doi:10.1016/S2213-2600(20)30079-5.

3. Bhatraju, P. K. et al. Covid-19 in Critically Ill Patients in the Seattle Region — Case Series. N. Engl. J. Med. 0, null (2020).

4. Stefan, N., Birkenfeld, A.L., Schulze, M.B. et al. Obesity and impaired metabolic health in patients with COVID-19. Nat Rev Endocrinol (2020). https://doi.org/10.1038/s41574-020-0364-6

5. Zhang, J. et al. Gender differences in the association between body mass index and health-related quality of life among adults:a cross-sectional study in Shandong, China. BMC Public Health 19, (2019).

6. Shereen, M. A., Khan, S., Kazmi, A., Bashir, N. & Siddique, R. COVID-19 infection: Origin, transmission, and characteristics of human coronaviruses. J. Adv. Res. 24, 91–98 (2020).

7. Muniyappa, R. & Gubbi, S. COVID-19 Pandemic, Corona Viruses, and Diabetes Mellitus. Am. J. Physiol.-Endocrinol. Metab. (2020) doi:10.1152/ajpendo.00124.2020.

8. Jia, X. et al. Two Things about COVID-19 Might Need Attention. (2020) doi:10.20944/preprints202002.0315.v1.

9. Hamming, I. et al. Tissue distribution of ACE2 protein, the functional receptor for SARS coronavirus. A first step in understanding SARS pathogenesis. J. Pathol. 203, 631–637 (2004).

10. Alti, D., Sambamurthy, C. & Kalangi, S. K. Emergence of Leptin in Infection and Immunity: Scope and Challenges in Vaccines Formulation. Front. Cell. Infect. Microbiol. 8, (2018).

11. Badawi, A. & Ryoo, S. G. Prevalence of comorbidities in the Middle East respiratory syndrome coronavirus (MERS-CoV): a systematic review and meta-analysis. Int. J. Infect. Dis. IJID Off. Publ. Int. Soc. Infect. Dis. 49, 129–133 (2016).

12. Kwong, J. C., Campitelli, M. A. & Rosella, L. C. Obesity and Respiratory Hospitalizations During Influenza Seasons in Ontario, Canada: A Cohort Study. Clin. Infect. Dis. Off. Publ. Infect. Dis. Soc. Am. 53, 413–421 (2011).

13. Honce, R. & Schultz-Cherry, S. Impact of Obesity on Influenza A Virus Pathogenesis, Immune Response, and Evolution. Front. Immunol. 10, (2019).

14. Yvan-Charvet, L. & Quignard-Boulangé, A. Role of adipose tissue rennin–angiotensin system in metabolic and inflammatory diseases associated with obesity. Kidney Int. 79, 162–168 (2011).

15. Ouchi, N., Parker, J. L., Lugus, J. J. & Walsh, K. Adipokines in inflammation and metabolic disease. Nat. Rev. Immunol. 11, 85–97 (2011).

16. Ibrahim, H. S., Froemming, G. R. A., Omar, E. & Singh, H. J. ACE2 activation by xanthenone prevents leptin-induced increases in blood pressure and proteinuria during pregnancy in Sprague-Dawley rats. Reprod. Toxicol. Elmsford N 49, 155–161 (2014).

17. Bellmeyer, A. et al. Leptin Resistance Protects Mice from Hyperoxia-induced Acute Lung Injury. Am. J. Respir. Crit. Care Med. 175, 587–594 (2007).

18. Yarandi, S. S., Hebbar, G., Sauer, C. G., Cole, C. R. & Ziegler, T. R. Diverse roles of leptin in the gastrointestinal tract: Modulation of motility, absorption, growth, and inflammation. Nutrition 27, 269–275 (2011).

19. Gane, S. B., Kelly, C. & Hopkins, C. Isolated sudden onset anosmia in COVID-19 infection. A novel syndrome? Rhinology (2020) doi:10.4193/Rhin20.114.

20. Savigner, A. et al. Modulation of Spontaneous and Odorant-Evoked Activity of Rat Olfactory Sensory Neurons by Two Anorectic Peptides, Insulin and Leptin. J. Neurophysiol. 101, 2898–2906 (2009).

21. Konstantinides, K. S., and S. Mechanisms Linking Leptin to Arterial and Venous Thrombosis: Potential Pharmacological Targets. Current Pharmaceutical Design vol. 20 635–640 http://www.eurekaselect.com/110202/article (2014).

22. Cohen, P. et al. Role for Stearoyl-CoA Desaturase-1 in Leptin-Mediated Weight Loss. Science 297, 240–243 (2002).

23. Zhang, J. et al. Therapeutic and triage strategies for 2019 novel coronavirus disease in fever clinics. Lancet Respir. Med. 8, e11-e12 (2020).

24. Maruna, P., Gürlich, R., Fraško, R. & Haluzík, M. Serum Leptin Levels in Septic Men Correlate Well with C-Reactive Protein (CRP) and TNF-alpha but not with BMI. 50, 6 (2001).

25. Ceddia, R. B., William, W. N. & Curi, R. Comparing effects of leptin and insulin on glucose metabolism in skeletal muscle: evidence for an effect of leptin on glucose uptake and decarboxylation. Int. J. Obes. 23, 75–82 (1999).

26. Ubags, N. D. J. et al. Hyperleptinemia is associated with impaired pulmonary host defense. JCI Insight 1,.

27. Szkudelska, K., Nogowski, L. & Szkudelski, T. The inhibitory effect of resveratrol on leptin secretion from rat adipocytes. Eur. J. Clin. Invest. 39, 899–905 (2009).

28. Bouloumié, A., Marumo, T., Lafontan, M. & Busse, R. Leptin induces oxidative stress in human endothelial cells. FASEB J. 13, 1231–1238 (1999).

29. Lian, N., Zhang, S., Huang, J., Lin, T. & Lin, Q. Resveratrol Attenuates Intermittent Hypoxia-Induced Lung Injury by Activating the Nrf2/ARE Pathway. Lung 198, 323–331 (2020).

30. Uchiyama T, Okajima F, Mogi C, Tobo A, Tomono S, Sato K. Alamandine reduces leptin expression through the c-Src/p38 MAP kinase pathway in adipose tissue. PlosOne 12, e0178769 (2017).

